# Datasheet for the IDHea Primary Care Screening Dataset: A Real-World Ocular Imaging Resource for Research

**DOI:** 10.1101/2025.04.29.25326705

**Authors:** Reena Chopra, Anya Guzman, Juho Uotila, John Bartolovich, Jonathan Liu, Anthony P. Khawaja, Pearse A. Keane, Matthew P. Lungren, Julia Coelho, Mary K. Durbin

## Abstract

**Purpose:** Ocular screening in primary care is increasingly recognized as a valuable opportunity to detect a wide range of eye conditions at an early stage, especially among individuals with systemic risk factors. The Primary Care Screening dataset is a large-scale, real-world collection of de-identified color fundus photographs (CFP) acquired during routine diabetic retinopathy (DR) screening visits in primary care clinics across the United States, and is available through the Institute for Digital Health (IDHea)—a secure research platform established by Topcon Healthcare, Inc.

**Methods:** The dataset includes CFP from individuals who participated in eye screening at 643 clinical sites in the United States. The majority of images were obtained using the TRC-NW400 (Topcon Corp., Tokyo, Japan), although other imaging devices were also used. Each image was graded by an eye care specialist using the International Clinical Diabetic Retinopathy (ICDR) grading system. Graders also recorded image quality and a wide range of other retinal findings. The dataset also includes patient-level demographics including age, sex, and 3-digit ZIP code, image metadata, along with model-generated annotations such as AutoMorph image quality, vascular metrics, and retinal pigment scores.

**Results:** As of March 2025, the dataset includes 427,182 CFP from 161,705 subjects, representing 372,528 eyes and 186,264 individual visits. Image quality was graded as Excellent or Good (218,139 eyes), Fair (109,127 eyes), or Unreadable (37,491 eyes). Most eyes (n = 293,391) had no diabetic retinopathy present (NDRP), while 15,514 had mild non-proliferative DR (NPDR), 10,856 moderate NPDR, 1,140 severe NPDR, and 1,644 proliferative DR. Diabetic macular edema was present in 4–34% of NPDR categories and 18% of proliferative DR cases. Additional findings included drusen or pigmentary changes (n = 9,649), glaucoma suspect (n = 4,257), and macular degeneration (n = 3,581).

**Conclusion:** The Primary Care Screening dataset represents one of the largest real-world collections of retinal images acquired in primary care settings. Its size, diversity, and detailed expert grading make it a valuable resource for research into automated screening, ocular disease prevalence, and AI model development. An independent Data Access and Governance committee oversees research applications to ensure responsible use. The dataset will be made available via the secure IDHea research platform. More information is available at IDHea.net.

## Introduction

The primary care clinic represents a critical opportunity for early detection of eye disease, particularly for individuals with systemic conditions such as diabetes. Without timely intervention, diabetic retinopathy (DR) can progress to severe visual impairment or blindness. In the United States, an estimated 16 million adults over the age of 40 are projected to be affected by DR by 2050, including 1.9 million with vision-threatening disease [1]. This underscores the urgent need for accessible, high-coverage screening programs to identify individuals at risk and ensure prompt treatment.

Although screening is effective in detecting both highrisk individuals and those with advanced disease, uptake remains suboptimal—only around 65% of individuals with diabetes who visited their primary care physician received a dilated eye exam in the prior year [2]. Integrating DR screening into primary care settings has been shown to substantially improve access to retinal exams and thereby prevent irreversible vision loss and improve quality of life [3].

To address this gap in screening coverage, *Healthcare from the Eye™ Screening*, an ocular screening initiative by THI, was launched in 2019. This program offers annual digital retinal photography to individuals with diabetes by embedding imaging devices within primary care practices, aiming to boost early detection and facilitate timely intervention. To date, over 160,000 individuals have been screened through this initiative, generating valuable real-world data to inform both clinical practice and research.

Recognizing the potential research value of this realworld dataset, THI has established the Institute of Digital Health (IDHea) to de-identify, curate, and securely host the dataset for research access. IDHea encompasses a data repository and secure workspace dedicated to research and innovation using ocular imaging and diagnostic data. The primary aim of IDHea is to transform population screening data into a research-ready format. The key IDHea attributes are:

- **Fast**: Streamlining data access requests to provide rapid access to a large repository of data to accelerate discovery and innovation.
- **Accessible**: Providing an intuitive and user-friendly platform that supports a diverse range of research initiatives, from exploratory analyses to machine learning model development, that can be accessed from anywhere, in the cloud.
- **Safe**: Employing industry-standard security protocols and infrastructure to provide the highest level of privacy and ensure compliance with health data protection regulations.
- **Transparent**: Maintaining a clear, open, and publicly available governance framework to build trust and ensure fair access to data. All data access requests are reviewed by an independent committee.

IDHea is designed to support fast, efficient, collaborative research by offering seamless access to high-quality, well-organized datasets. Researchers across academic and industrial sectors can request access through a transparent governance process overseen by an independent Data Access and Governance Committee (DAG). The DAG is responsible for reviewing and approving data access requests, ensuring that data usage aligns with ethical guidelines and the broader goal of advancing ophthalmic research to improve patient outcomes.

This datasheet provides an overview of the IDHea Primary Care Screening dataset, outlining the collection, composition, and characteristics, and follows guidelines set out by Rostamzadeh et al. 2022 [4].

## Datasheet

### Dataset composition

The IDHea Primary Care Screening dataset is routinely collected data through the *Healthcare from the Eye Screening* program. The program has been implemented in 643 physician offices across 36 states in the US, since January 2019 (Figure 1). ZIP codes of each office, stripped to the first 3 digits, are available for analysis. Primary care providers in this program follow American Diabetes Association (ADA) guidelines for diabetic eye screening [5]. These recommend that individuals with type 2 diabetes begin annual eye exams at the time of diagnosis, while those with type 1 diabetes begin annual screening five years after initial diagnosis—typically during adolescence or early adulthood. All individuals with diabetes should be screened annually unless they have already been diagnosed with more than mild DR, in which case ongoing management should occur under the care of an eye care professional. If no record of a recent eye exam is available, the primary care provider should proceed with screening.

**Fig. 1.**
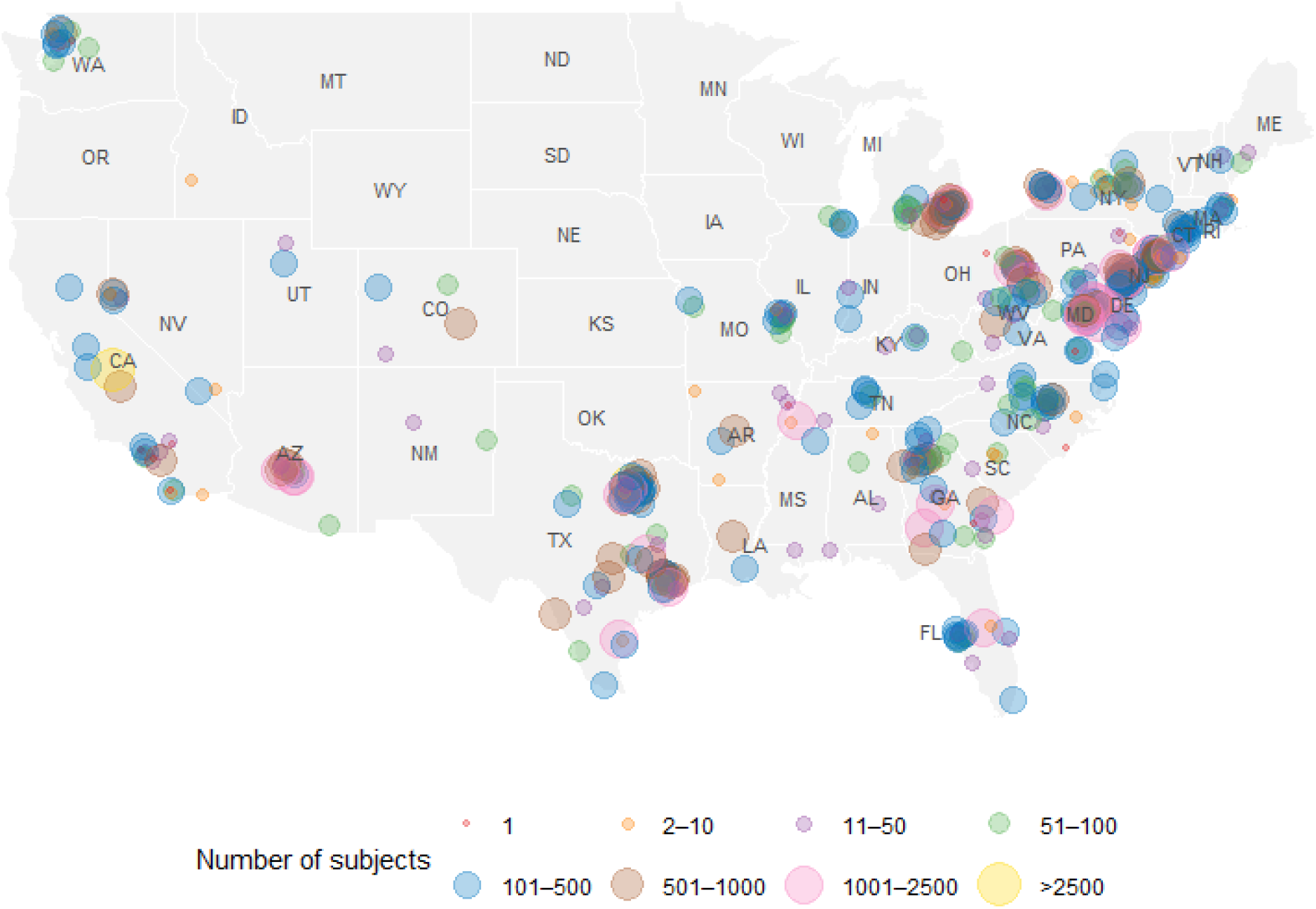
Geographic distribution of subjects in the Primary Care Screening dataset, aggregated by ZIP code. Dot size and color represent the number of unique subjects per ZIP code. A total of 145 patients were missing ZIP codes. ZIP code with the first 3 digits “098” was associated with 248 subjects and could not be mapped due to the absence of geographic coordinates in the reference ZIP code database; this ZIP is reserved for military or diplomatic addresses and does not correspond to a standard geographic location.

At each visit, color fundus photographs (CFP) of both eyes are captured (where applicable), using either a tabletop non-mydriatic retinal camera (TRC-NW400 or NW500, Topcon Corporation, Tokyo, Japan) or a handheld device (Signal, Optomed Plc., Oulu, Finland; or Horus Scope, Medimaging Integrated Solution Inc., Hsinchu, Taiwan). All sites used a retinal camera that was connected to a unified instance of Topcon Harmony®, a clinical image and data management software platform. Each set of images for a given visit was graded by a single expert—either an optometrist or ophthalmologist licensed to practice within the state for which they read. Grading was performed using a structured report form within the Harmony software that included fields for image quality, DR severity, and other retinal findings. Report versions are provided in the dataset. In versions 1–4 of the grading form, image quality was categorized as “Excellent,” “Good,” “Fair,” or “Unreadable.” From version 5 onward, the “Excellent” category was removed, with available options limited to “Good,” “Fair,” and “Unreadable”. Images that were deemed readable were then graded for DR severity using the International Classification of Diabetic Retinopathy (ICDR) system: no DR present (NDRP), mild non-proliferative diabetic retinopathy (NPDR), moderate NPDR, or severe NPDR, proliferative DR, and diabetic macular edema (DME). While the ICDR provided a standardized framework, graders also applied clinical judgment when assigning grades.

In addition to DR grading, the form allowed documentation of other notable retinal findings, including choroidal nevus, cotton wool spots, drusen, epiretinal membrane (ERM), glaucoma suspicion, increased cup-to**-**disc ratio, macular degeneration, and vascular tortuosity. A free-text field was available to capture additional retinal findings not covered by the structured proforma.

A corresponding screening outcome was assigned at the visit level, including options such as re-screening if the photo was unreadable, no referral required, or referral to an eye care specialist with a level of urgency (‘Normal’ if routine and ‘Urgent’ otherwise).

### Ethics

Data sharing agreements were established with each participating primary care site, permitting the use of de-identified data for research purposes. In instances where the site did not have the technical capabilities for de-identifying the data on their own, a site executed a Health Insurance Portability and Accountability Act (HIPAA) Business Associate Agreement (BAA) to allow secure transfer of identifiable information for de-identification by a technology team. Only after the data had been de-identified, in accordance with HIPAA Privacy Rule’s Safe Harbor method (45 C.F.R. § 164.514(b)(2)), were datasets made accessible to the study team for research use. Under HIPAA, once data have been de-identified according to this standard, they are no longer considered protected health information (PHI) and may be used or shared for research purposes without patient authorization. De-identified datasets were securely transferred and stored in accordance with the security and data protection policies applicable to the data platform. This approach ensures compliance with ethical and regulatory standards for the secondary use of clinical data in research while protecting patient privacy and enabling broader learning from real-world healthcare delivery.

### Data collection and de-identification

The data were collected retrospectively from primary care sites participating in the program.

Prior to any research use, the data, including imaging and grading data, were de-identified in accordance with the HIPAA Safe Harbor requirements, which included the removal of 18 direct and quasi-identifiers. The de-identification process was performed using a standardized process: each patient and visit was assigned a pseudonym identifier, by cryptographically hashing a salted internal database identifier. For eyes with multiple images captured during the same visit, the images share the same de-identified visit identifier and include a numeric suffix in the filename to indicate the capture order. De-identified subject identifiers remain consistent across multiple visits. Ages above 90 years are recorded as 90 to reduce the risk of re-identification.

### Pre-processing and labeling

All images are stored in JPEG format to ensure portability and avoid sensitive metadata leakage. Images had a pixel width and height of 1956 x 1934 for TRC-NW400, 2676 x 2676 for NW500, 2368 x 1776 for Signal, 2560 x 1920 for Horus Scope.

Grading data were curated and structured into standardized tables at both the visit and eye level. DR grades and other retinal findings were standardized using a consistent encoding scheme: 1 for present, 0 for absent, and −1 for eyes that had unreadable images or had missing grades.

CFP were also processed by several artificial intelligence (AI) models, with their outputs included to support research without requiring users to implement their own image analysis pipelines. AutoMorph, an AI-based model for CFP, is included to assess image quality and extract features related to the optic disc and blood vessels [6]. Retinal pigmentation score was applied to CFP deemed good or usable quality by AutoMorph [7].

To facilitate AI model validation, a standardized and representative test subset was held out using propensity score matching (PSM), enabling consistent benchmarking and comparison of algorithm performance across studies. This subset, comprising approximately 10% of individuals from the whole dataset, was selected to reflect the broader population in terms of key covariates: age, gender, laterality, retinal camera, and DR grade. PSM was applied to balance these characteristics, ensuring that the selected test set is representative of the full dataset while maintaining the real-world diversity of routine optometric imaging. Access to this subset is limited and requires submission of a formal request through the same governance and review process as the full dataset.

### Dataset characteristics

Summary statistics are reported prior to separating the test subset. As of March 2025, the dataset includes 427,182 CFP from 161,705 subjects, representing 372,528 eyes and individual 186,264 visits. The mean age at image acquisition was 59.6 years (SD = 14.6) (Figure 2). 908 (0.5%) were reported as age 0-5 years at a given visit, which are likely to be data entry errors. 46.9% of subjects were reported as female, 52.7% as male, and 0.4% other or not reported. Follow-up data is available for 19,775 individuals who had more than one visit. Of these, 15,834 had two visits, 3,209 had three visits, 640 had four visits, and 92 had five or more visits. Note that additional visits may occur if images were Unreadable and required repeat imaging. The mean age difference between visits was 1.35 years (SD = 0.95).

**Fig. 2.**
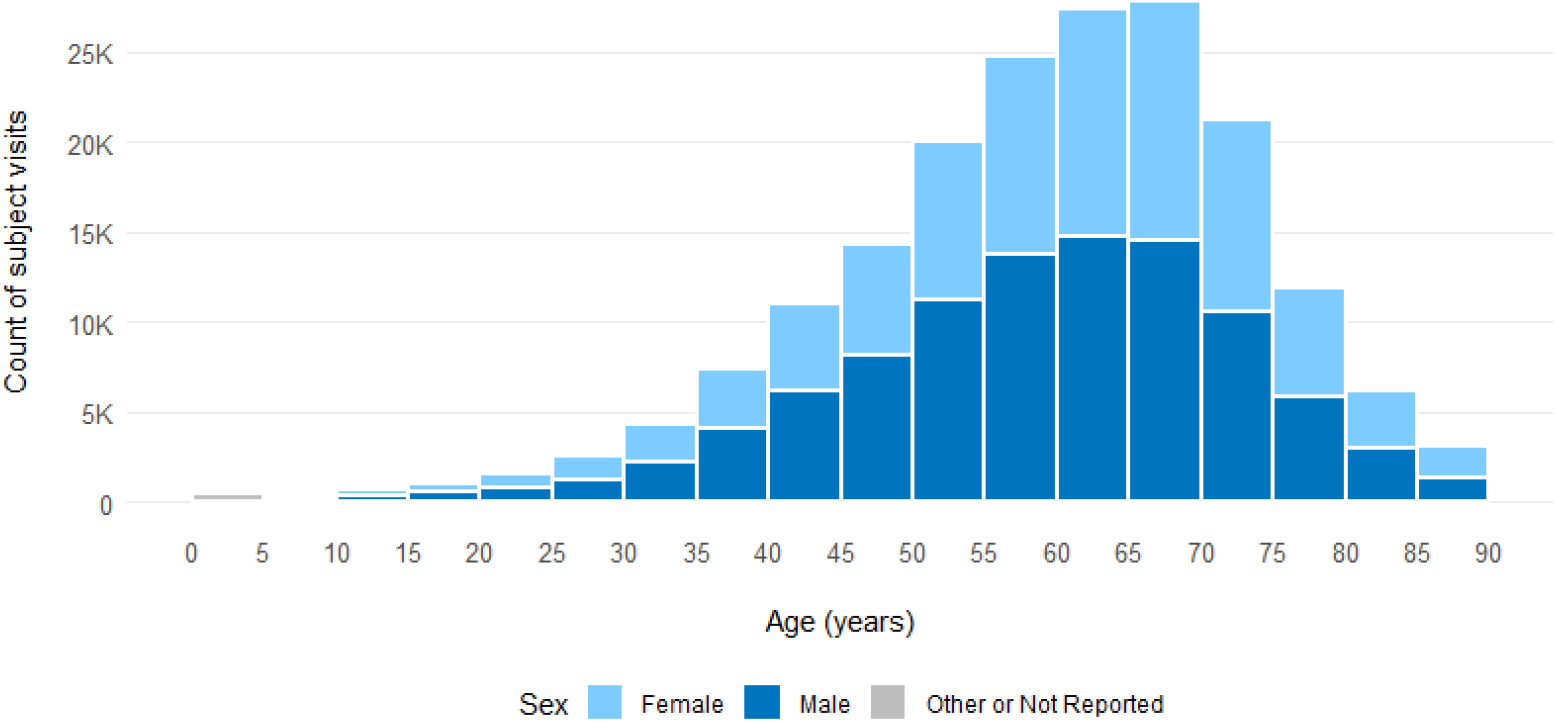
Distribution of age at visit, stacked by sex.

The majority of CFP were captured using the TRCNW400, which accounted for 395,050 images (91.3%) from 351,446 eyes. The Signal device captured 28,912 images (6.7%) from 18,080 eyes, followed by the NW500 with 2,163 images (0.5%) from 2,122 eyes, and the Horus Scope with 1,057 images (0.2%) from 880 eyes.

Of the eyes graded for image quality, 29,248 (7.9%) were assessed as “Excellent,” 188,891 (50.7%) as “Good,” 109,127 (29.3%) as “Fair,” and 37,491 (10.0%) as “Unreadable” by the expert grader. An additional 7,772 (2.1%) eyes had no recorded image quality grade; however, 1,308 of these still received a DR grade.

Most eyes (n = 293,391, 78.8%) were graded with NDRP, while 15,514 (4.2%) had mild non-NPDR, 10,856 (2.9%) had moderate NPDR, 1,140 (0.3%) had severe NPDR, and 1,644 (0.4%) had proliferative DR. Among eyes with mild NPDR, 4% also had diabetic macular edema (DME), increasing to 23% in moderate NPDR, 34% in severe NPDR, and 18% in proliferative DR (Figure 3). 37,851 (10.2%) eyes were not given a DR grade, the majority of which were deemed Unreadable.

**Fig. 3.**
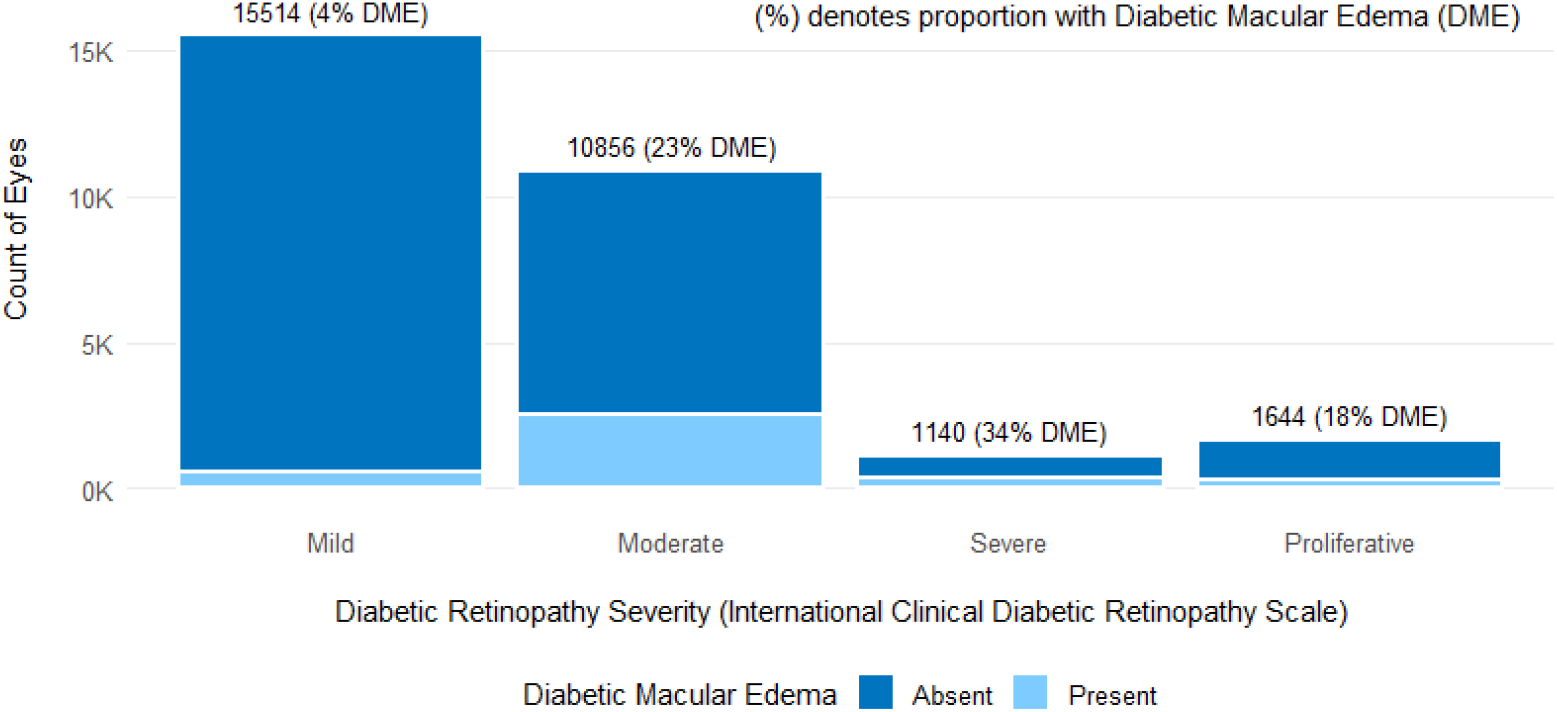
Stacked bar chart showing the prevalence of diabetic macular edema (DME) by diabetic retinopathy (DR) severity. DR severity was graded using the International Clinical Diabetic Retinopathy Scale.

The most frequently observed ancillary ocular findings were drusen or pigmentary changes (n eyes = 9,649), increased cup-to-disc ratio (n = 7,070), and glaucoma suspect (n = 4,257) (Figure 4). Other common findings included macular degeneration (n = 3,581), epiretinal membrane (n = 3,218), and vascular tortuosity (n = 2,313). Less prevalent findings were choroidal nevus (n = 1,441) and cotton wool spots (n = 589) (Figure 4).

**Fig. 4.**
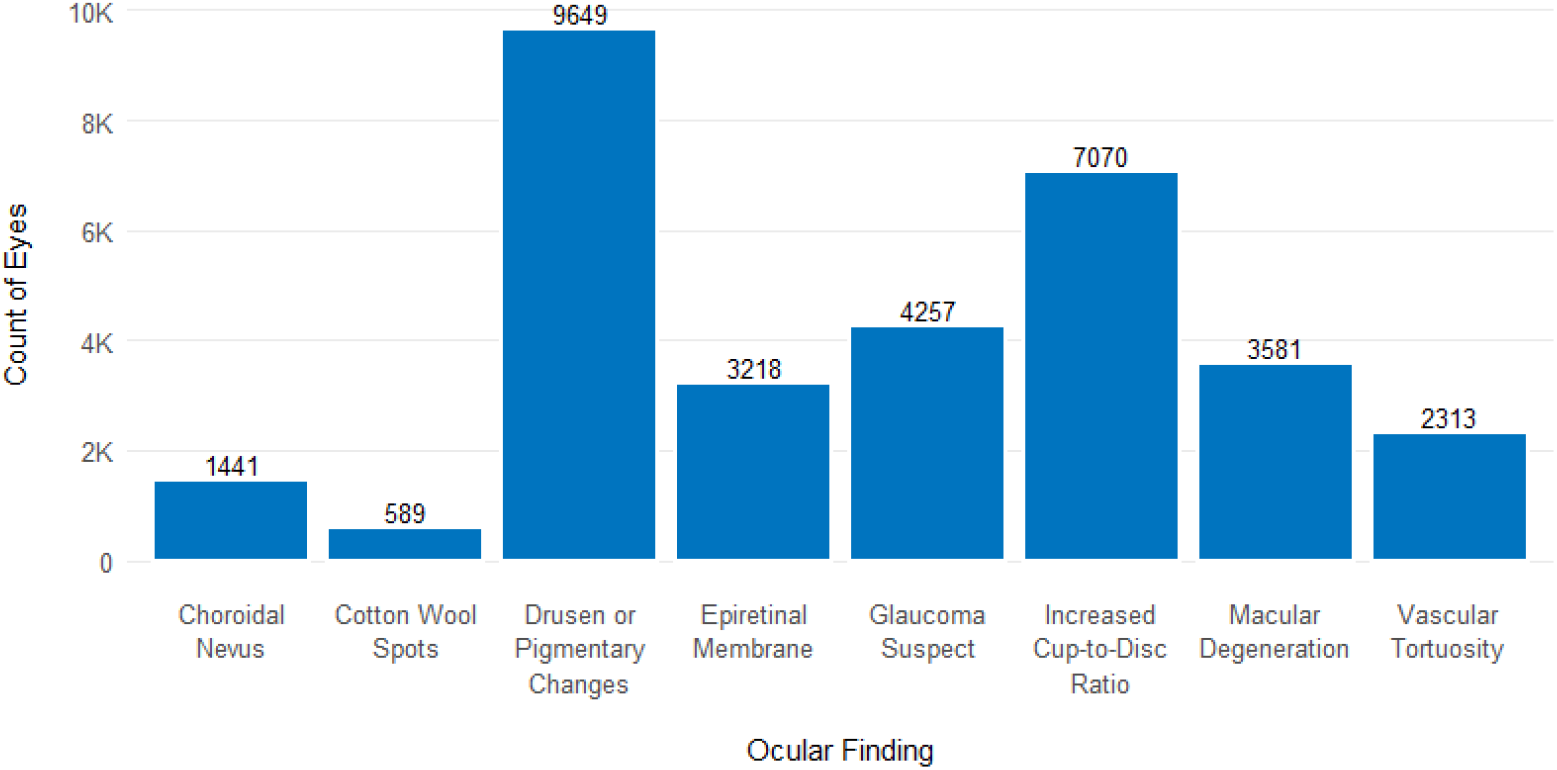
Number of eyes graded with other ocular findings. Multiple findings can be recorded per eye.

AutoMorph image quality assessment classified 37.2% of images as “good” quality, 19.2% as “usable” quality, and 43.6% of images as “bad” quality. In contrast, expert graders marked only 10.0% of eyes as unreadable.

### Dataset versioning

This datasheet summarizes the first version of the dataset release, comprising data collected from January 2019 up to March 2025. There are plans to release new major versions of the dataset periodically with additional data collected from new sites and/or existing sites. For existing sites, new records will be matched to previously collected data using consistent de-identification methods, ensuring that each unique subject retains the same identifier across versions. Sub-versions will be released on an ongoing basis which will include additions such as labels to an existing version.

### Data access and governance

The IDHea Primary Care Screening dataset is available to researchers across academic and industrial sectors through a structured application process designed to ensure ethical, secure, and responsible data use. Applicants must submit their research proposal outlining their objectives, methodologies, and anticipated outcomes for evaluation.

The IDHea Data Access and Governance committee, an independent body, is responsible for reviewing data access requests and ensuring alignment with ethical principles and governance policies. Comprising experts in clinical practice, research, and technology, the committee provides multidisciplinary oversight to safeguard responsible data use. All members serve in a personal capacity, acting independently of their institutional or commercial affiliations to ensure impartial and unbiased decision-making.

The committee establishes criteria for evaluating research and development applications, focusing on adherence to IDHea’s governance policies, ethical standards, and alignment with previously approved or precedent use cases. Each application undergoes a structured review to confirm eligibility, including verification of the applicant’s identity and affiliation and the suitability of the proposed use. Similar or identical use cases may be approved for multiple applicants, and assessments are grounded in policy and precedent rather than scientific evaluation. This process ensures consistency, fairness, and responsible use of the dataset.

Transparency is a fundamental principle of the committee’s operations. Every data access request is reviewed against predefined governance policies, with documented findings ensuring clear accountability. Where applications are approved and data access is granted, the project title and name of the lead researcher will be published online at IDHea.net to uphold integrity and trust in the governance process and research use.

A tiered access model with discounts for academic users supports broad participation in research while ensuring the sustainability of the data infrastructure. Access to additional resources, such as expert annotations or AI model-generated labels, will incur fees for both academic and commercial users, reflecting the added cost and effort involved in producing and maintaining these enriched datasets.

### Secure cloud computing environment

The dataset is hosted within a secure workspace on Microsoft Azure Databricks [8], ensuring a controlled environment with no data egress while providing integrated tools for data analysis, machine learning, and collaborative development (Figure 5). Microsoft Azure Databricks is HITRUST CSF certified and is designed to support compliance with standards like HIPAA, GDPR, and prevailing industry standards [9]. Researchers benefit from interactive notebooks, SQL-based querying, custom dashboards, and scalable compute clusters, all within a unified platform designed to streamline large-scale data science. While raw data remains within the IDHea secure workspace, users can export their trained model weights enabling them to apply their models in external workflows. To further protect data integrity, agreements are in place with researchers to ensure that any exports do not include elements that would allow for reconstruction or replication of the original dataset.

**Fig. 5.**
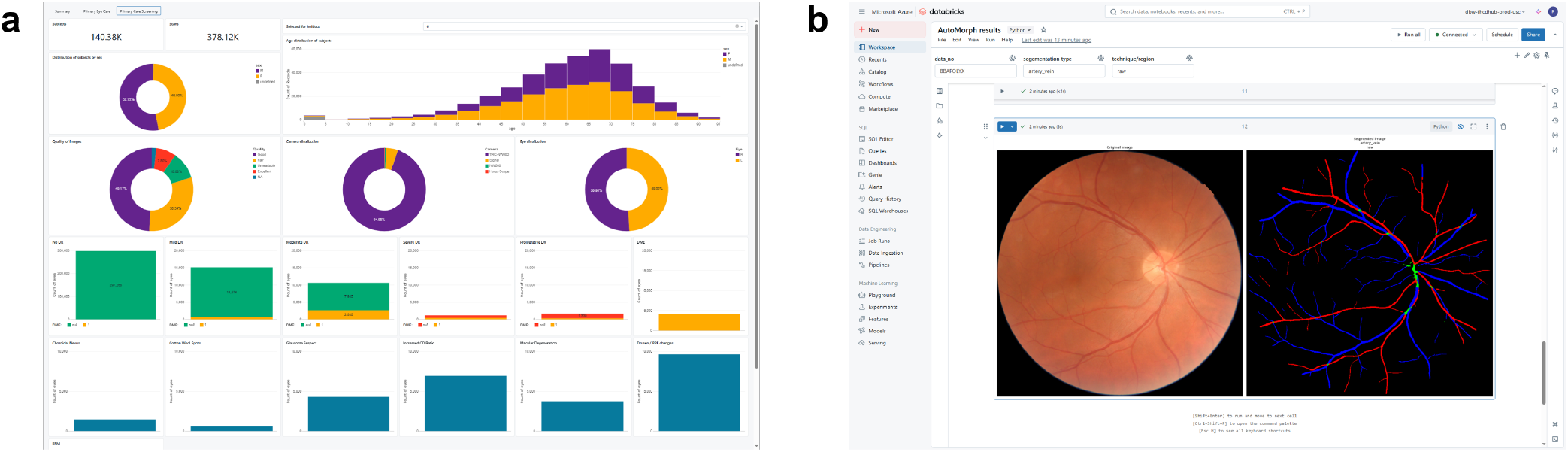
Microsoft Azure Databricks workspace. a) Interactive dashboards for data exploration. b) Reading color fundus photographs and associated AutoMorph segmentation outputs.

To streamline research workflows, example notebooks are provided within the secure workspace, demonstrating how to read files, pre-process images, and perform basic machine learning development and evaluation. These notebooks offer step-by-step guidance for researchers, facilitating efficient data exploration and model development without requiring extensive setup.

Additionally, a cloud-based computing infrastructure enables access to GPUs for high-performance computing, supporting advanced computational tasks and democratizing access to powerful resources. In addition, cost management tools are available, enabling researchers to monitor and optimize compute usage for sustainable, cost-effective access to resources.

### Strengths and limitations

The IDHea Primary Care Screening dataset offers several notable strengths. It comprises over 427,182 color fundus photographs from 161,705 individuals across 643 primary care offices in 36 U.S. states, making it one of the largest and most geographically diverse collections of retinal images captured in routine primary care. The dataset reflects real-world variability in imaging conditions, patient demographics, and device usage, enhancing its relevance for studying diagnostic performance and generalizability. The data are hosted in a secure, cloud-based research environment equipped with tools for scalable analysis, model development, and benchmarking.

Within this environment, images are stored in a standardized, research-ready format, where they are linked to expert-curated diabetic retinopathy grades and image quality scores, enabling efficient development and validation of automated analysis tools. A held-out, propensity score–matched test subset is also available to enable standardized comparisons across studies.

Several limitations should be acknowledged. The dataset is designed for screening and does not include comprehensive medical records, systemic health data, or follow-up outcomes beyond image-based assessments. Most individuals have only a single recorded screening visit, limiting opportunities for longitudinal analysis. While follow-up data are available for a subset of patients, this may introduce bias in studies of disease progression. A small number of implausible entries, such as patients recorded as under age five at the time of imaging, suggest minor data entry errors given that screening for type 1 diabetes typically starts five years after diagnosis. Each image was graded by a single eye care expert, without adjudication or consensus grading, which may contribute to variability in DR severity labels. Prior studies have shown only moderate agreement among experts grading diabetic retinopathy, with inter-grader kappa values often ranging from 0.40 to 0.65 depending on grading experience and methodology [10, 11].

Lastly, although most images were acquired using a single tabletop camera model, several different devices were used across sites, contributing to variability in image quality and capture conditions. Additionally, variability in device operators, such as differences in training, experience, and imaging technique, may also impact image quality and consistency across the dataset. This is reflected in AutoMorph assessments, which classified 43.6% of images as “bad” quality, despite only 10% of eyes being marked as unreadable by expert graders. This discrepancy likely reflects real-world screening conditions, where images may fall below algorithmic quality thresholds yet remain clinically usable for detecting eye disease. It underscores the distinction between technical image quality and clinical interpretability in real-world settings, and the importance of incorporating both expert and AI-based quality assessments in future research.

## Summary

The IDHea Primary Care Screening dataset provides a large-scale, labeled ocular imaging resource that supports research into diabetic retinopathy and other retinal pathologies within real-world primary care settings. Its breadth and structure enable population health studies, the development of clinical decision support systems, and the validation of AI-driven diagnostic tools.

This dataset builds upon existing real-world ocular imaging resources that have significantly advanced large-scale eye research. The Illinois Ophthalmic Database Atlas (I-ODA) offers a multi-modal, longitudinal dataset that enables detailed clinical studies and disease progression analysis [12]. INSIGHT, a UK-based health data research hub, aggregates anonymized eye scans including a dataset from diabetic retinopathy screening programs, to support AI-driven disease detection and healthcare innovation [13–15]. The UK Biobank provides extensive ocular imaging data linked to genetic and health records, allowing for large-scale population health studies [16]. Widely used open-access diabetic retinopathy datasets such as EyePACS [17], APTOS 2019 [18], and MESSI-

DOR [19] have played a pivotal role in the development of deep learning algorithms by providing large-scale labeled fundus photographs for model training and evaluation. More recently, the DDR dataset has provided a valuable resource for DR research across hospital systems in China, offering diverse patient data and lesion-level annotations [20]. Finally, the AI-READI consortium have generated and publicly released a dataset for the study of type 2 diabetes, comprising over 1,000 participants and 15 data modalities including several OCT devices, designed specifically to support AI research [21].

The IDHea dataset complements these efforts by focusing on primary care settings—an environment underrepresented in existing datasets. Its grounding in everyday clinical workflows makes it especially valuable for evaluating how AI tools and clinical decision support systems might perform when deployed in real-world frontline healthcare. Expanding access to diverse, representative imaging data is essential to ensuring that innovations in screening not only advance scientific discovery, but also improve health equity by reaching patients who are often underserved by traditional specialty care pathway. This forms part of a broader suite of research datasets under the IDHea umbrella, which includes the Primary Eye Care dataset, offering optical coherence tomography and ophthalmic photography from optometric settings, and the Multimodal Healthy Eyes dataset, comprising OCT, CFP, visual fields, and other diagnostic information from a prospective clinical study. Together, these datasets provide a valuable foundation for AI model development, validation, and deployment in diverse real-world contexts.

With transparent governance and secure cloud access, researchers can analyze data within a controlled environment while exporting trained model weights for external application. Regular updates will further expand the dataset’s scope, enhancing its utility for AI development, epidemiological studies, and clinical research.

Data access can be requested by submitting an application at IDHea.net.

## Acknowledgements

We gratefully acknowledge the following individuals for their thoughtful feedback and review of the manuscript: Marco Miranda, Carole McCallum, Kerry Goetz, Jacqueline Armani, Ray Everett, Yvonne Bentley, and Ali Tafreshi.

## Declarations

### Data availability

Applications for data access can be submitted via IDHea.net. A sample dataset is freely available for download following registration.

### Code availability

Example notebooks written in Python are available within the Databricks environment.

### Author contribution

R.C., J.B., J.L., J.C., M.K.D. conceptualized the project. R.C., A.G., J.U., J.B., J.L., M.K.D. collected, curated and processed the data. R.C., A.G. performed the analysis and visualization. R.C. drafted the manuscript. All authors reviewed and edited the manuscript. J.C. and M.K.D. supervised the project.

